# MANTIS at #SMM4H 2023: Leveraging Hybrid and Ensemble Models for Detection of Social Anxiety Disorder on Reddit

**DOI:** 10.1101/2023.12.05.23299439

**Authors:** Sourabh Zanwar, Daniel Wiechmann, Yu Qiao, Elma Kerz

## Abstract

This paper presents our system employed for the Social Media Mining for Health 2023 Shared Task 4: Binary classification of English Reddit posts self-reporting a social anxiety disorder diagnosis. We systematically investigate and contrast the efficacy of hybrid and ensemble models that harness specialized medical domain-adapted transformers in conjunction with BiLSTM neural networks. The evaluation results outline that our best performing model obtained 89.31% F1 on the validation set and 83.76% F1 on the test set.

## 1 Introduction

According to the Anxiety & Depression Association of America^1^, anxiety disorders rank as the most prevalent mental illnesses in the United States. An estimated 40 million adults, constituting 19.1% of the population aged 18 and above, grapple with these conditions annually. This challenge is compounded by the scarcity of accessible mental health care services and the frequent occurrence of misdiagnoses, often causing individuals to unknowingly endure these disorders (Kasper, 2006).

Natural Language Processing in combination with Machine Learning is increasingly recognized as having transformative potential to support healthcare professionals and stakeholders in the early detection, treatment and prevention of mental disorders (Zhang et al., 2022). In this paper, we report on our participation in The Social Media Mining for Health Applications (#SMM4H) 2023 workshop, which aims to promote automated methods for mining social media data for health informatics. We chose to participate in the Shared Task 4 competition, which was to improve social anxiety detection in Reddit posts (Klein et al., 2023). We approached this task by developing hybrid and ensemble models combining domain-matched transformers with Bidirectional Long Short-Term Memory (BiLSTM) networks trained on a comprehensive set of engineered linguistic features. This set encompasses measures of morpho-syntactic complexity, lexical sophistication/diversity, readability, stylistics measures (register-specific ngram frequencies) and sentiment/emotion lexicons.

## 2 Data

The data for Task 4 consisted of 8117 Reddit posts written by users aged between 12 and 25 years. These data were split into training (75%), validation (8.4%), and testing sets (16.6%).

In preparation for model training, all texts were subjected to preprocessing procedures including eliminating HTML, URLs, excessive spaces, and emojis from the text, as well as rectifying inconsistent punctuation.

## 3 System Description

Our systems leveraged three domain-adapted Transformer-based pretrained language models (PLM), a BiLSTM trained on engineered features and their combination forming into hybrid and ensemble models. Domain-adapted pretrained language models include: (1) PsychBERT (Vajre et al., 2021) (2) Mental RoBERTa (Ji et al., 2022) and (3) Clinical BERT (Alsentzer et al., 2019). All PLMs were obtained from the Huggingface (Wolf et al., 2020), choosing the uncased, where applicable, base versions. We constructed a BiLSTM trained on 168 features that fall into six categories. All measurements of these features were obtained using a system that employs a sliding window technique to compute sentence-level measurements. The BiLSTM model is formulated as:

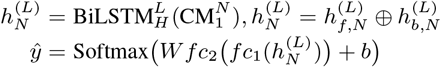

where *fc*_*i*_(*x*) = ReLU(*W*_*i*_*x* + *b*_*i*_), 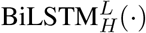 is a *L* layer BiLSTM with hidden size of *H*. 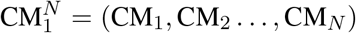, where CM_*i*_ represents the linguistic features for the *i*th sentence of a post consisting of *N* sentences. The last hidden representation of the last layer in forward and backward directions are denoted by 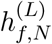 and 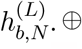 denotes the concatenation operator.

The hybrid model combines a Mental RoBERTa model with above BiLSTM.

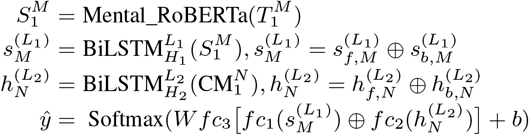

where 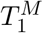 is the sequence of tokens from a post.

We constructed three distinct ensemble models using the stacking method: ensemble model (1) composed of instances from the hybrid model, which emerged as the most accurate base model (M6), (2) combining hybrid models with fine-tuned PsychBERT models (M7), and (3) consisting of Mental RoBERTa models, PsychBERT models, and BiLSTM models (M8). The resulting models represent homogeneous ensemble (HOE), intermediate and heterogeneous ensemble (HEE) approaches (Ganaie et al., 2022). As meta-learners, Support Vector Classifer, Logistic Regression, Gradient Boosting and Ridge Regression and XGBoost were used.

Further details are provided in the supplementary material (https://shorturl.at/epuF3).

## 4 Results and Evaluation

The results of our models on the validation set and test set are presented in Table 1. The Mental RoBERTa model achieved the highest performance (F1=86.59%) among the PLMs, outperforming the PsychBERT and ClinicalBERT models by 4% and 14.73%, respectively. This finding indicates that the detection of anxiety on Reddit sees a marked improvement from pretraining the PLM on mental health-related subreddits, as opposed to pretraining on clinical text. The hybrid models consistently outperformed the standalone PLM across all model iterations, yielding an average increase in F1 scores of 0.3%. The use of model stacking enhanced classification outcomes with performance boosts ranging between 1.86% and 2.44% in F1 score. The highest balanced classification score was achieved by the HEE model (M8). A variant of this model using ridge regression as a meta-learner (M12) achieved the best performance on the test set (F1 = 83.76%, mean_all teams_ = 79.3%, median_all teams_ = 82.4%). The HOE model (M6) achieved the second-highest performance and the best precision among all models examined. This suggests that both ensemble approaches can produce beneficial, albeit distinct, impacts on the detection of social anxiety disorder.

**Table 1:** Results on the validation set (top) and test set (bottom). For each ensemble model, we report results of the best performing meta-learner.

| Detection model | F1 | P | R |
| --- | --- | --- | --- |
| *Pretrained Language Models* |  |  |  |
| M1: Mental RoBERTa | 86.59 | 81.85 | 91.91 |
| M2: PsychBERT | 82.59 | 79.46 | 85.98 |
| M3: ClinicalBERT | 71.86 | 69.36 | 74.55 |
| M4: BiLSTM | 59.01 | 60.03 | 58.92 |
| M5: Hybrid Model | 86.87 | 83.30 | 90.76 |
| *Ensemble Models* |  |  |  |
| M6: HOE M5 (GB) | 88.80 | 85.57 | 92.28 |
| M7: HEE M5+M2 (GB) | 88.73 | 84.90 | 92.92 |
| M8: HEE M1+M2+M4 (GB) | 89.31 | 85.02 | 94.06 |
| *Test set* |  |  |  |
| M6: HOE: M5 (GB) | 83.63 | **81.07** | 86.35 |
| M12: HEE M1+M2+M4 (Ridge) | **83.76** | 80.6 | **87.17** |

## Data Availability

This paper has been peer-reviewed and accepted for presentation at the #SMM4H 2023 Workshop. The dataset used in this paper is a part of #SSM4H 2023 shared task. More information can be found at: https://healthlanguageprocessing.org/smm4h-2023/

## Footnotes

1 https://adaa.org/understanding-anxiety/facts-statistics

